# High rates of SARS-CoV-2 infection in pregnant Ugandan women and association with stunting in infancy

**DOI:** 10.1101/2023.06.16.23291450

**Authors:** Karen B. Jacobson, Katharina Röltgen, Brandon Lam, Patience Nayebare, Abel Kakuru, Jimmy Kizza, Miriam Aguti, Felistas Nankya, Jessica Briggs, Saki Takahashi, Bryan Greenhouse, Isabel Rodriguez-Barraquer, Kattria van der Ploeg, Jacob N. Wohlstadter, George B. Sigal, Michelle E Roh, Joaniter I. Nankabirwa, Gloria Cuu, Stephanie L. Gaw, Philip J. Rosenthal, Moses R. Kamya, Isaac Ssewanyana, Grant Dorsey, Scott D. Boyd, Prasanna Jagannathan

## Abstract

**Background:** SARS-CoV-2 has been well studied in resource-rich areas but many questions remain about effects of infection in African populations, particularly in vulnerable groups such as pregnant women.

**Methods:** We describe SARS-CoV-2 immunoglobulin (Ig) G and IgM antibody responses and clinical outcomes in mother-infant dyads enrolled in malaria chemoprevention trials in Uganda.

**Results:** From December 2020 to February 2022, among 400 unvaccinated pregnant women, serologic assessments revealed that 128 (32%) were seronegative for anti-SARS-CoV-2 IgG and IgM at enrollment and delivery, 80 (20%) were infected either prior to or early in pregnancy, and 192 (48%) were infected or re-infected with SARS-CoV-2 during pregnancy. We observed preferential binding of plasma IgG to Wuhan-Hu-1-like antigens in individuals seroconverting up to early 2021, and to Delta variant antigens in a subset of individuals in mid-2021. Breadth of IgG binding to all variants improved over time. No participants experienced severe respiratory illness during the study. SARS-CoV-2 infection in early pregnancy was associated with lower median length-for-age Z-score at age 3 months compared with no infection or late pregnancy infection (- 1.54 versus −0.37 and −0.51, p=0.009).

**Conclusion:** Pregnant Ugandan women experienced high levels of SARS-CoV-2 infection without severe respiratory illness. Variant-specific serology testing demonstrated evidence of antibody affinity maturation at the population level. Early gestational SARS-CoV-2 infection was associated with shorter stature in early infancy.

**Funding:** This work was supported by: Stanford MCHRI/Stephen Bechtel Endowed Fellowship in Pediatric Translational Medicine (KJ), Swiss National Science Foundation PRIMA grant PR00P3_208580 (KR), the Bill and Melinda Gates Foundation, and NIAID (T32-AI052073, U01- AI141308, U01-AI155325).

## Research in Context

### Evidence before this study

Africa has experienced fewer COVID-19 cases and deaths compared to the Americas and Europe. Possible reasons for this include a younger population with fewer underlying health conditions, underreporting of cases, and potential differences in immune responses.

Retrospective serological studies can help understand the spread of the virus in Africa despite lack of available testing in real time. Large cohort studies and meta-analyses from varied populations have shown that SARS-CoV-2 infection during pregnancy causes more severe disease in mothers and may lead to adverse birth outcomes including preterm birth, but data from regions of Africa where malaria in pregnancy is also a concern is extremely limited.

### Added value of this study

This study describes one of the largest SARS-CoV-2 in pregnancy cohorts from malaria-endemic Africa with infant follow up to date. We compare pre-pandemic SARS-CoV-2 antibody responses in Ugandan and U.S. adults, showing that Ugandans did not have higher pre-pandemic exposure to human coronaviruses. We characterize SARS-Co-2 infections in a Ugandan malaria pregnancy cohort, in which no severe respiratory illness occurred despite high infection rates in 2021. We also investigate the impact of SARS-CoV-2 infection in pregnancy on birth and infant outcomes in Uganda, and report an association between SARS-CoV-2 in early pregnancy and infant stunting. Using variant-specific SARS-CoV-2 serology testing, we demonstrate evidence of antibody affinity maturation at the population level.

### Implications of all the available evidence

Although SCV2 in pregnancy may not result in significant illness in many pregnant women living in malaria endemic African settings, it may result in long-term sequelae in infants.

## Background

In contrast to soaring coronavirus disease 2019 (COVID-19) case numbers during pandemic waves in the Americas and Europe, large parts of Africa have reported relatively low numbers of COVID-19-related morbidity and death (1, 2). Possible explanations for this observation include a younger population age structure with lower rates of comorbidities associated with more severe COVID-19 (3), underreporting of the true disease burden in Africa (4), and potential differences in immunological background of populations, including cross-protective immunity from prior exposure to endemic coronaviruses or “trained” immunity from other pathogen exposures in African populations (5, 6). Studies investigating immune responses to severe acute respiratory syndrome coronavirus 2 (SARS-CoV-2) infection in African compared to U.S. or European populations and their effects in pregnancy are limited.

Testing pre-pandemic plasma samples for antibodies to SARS-CoV-2, as well as to SARS-CoV- 1, Middle East Respiratory Syndrome (MERS), and other endemic human coronaviruses (HCoVs) such as HCoV-229E, HCoV-HKU1, HCoV-NL63, HCoV-OC43, provides information on pre-existing cross-reactive antibody responses that may protect against severe COVID-19 in certain populations. Given the lack of widespread access to SARS-CoV-2 reverse-transcription polymerase chain reaction (RT-PCR) or rapid antigen testing in many African countries during the pandemic (7), serologic testing of banked plasma specimens for the presence of anti-SARS-CoV- 2 antibodies is a useful tool for understanding the epidemiology of SARS-CoV-2 infection in Africa (8). The most common targets for serological assays are the Spike (including the receptor-binding domain, RBD) and Nucleocapsid (N) proteins, as most individuals infected with SARS-CoV-2 develop antibodies to these antigens (9). RBD mediates viral attachment and is a dominant target of anti-SARS-CoV-2 neutralizing antibodies. During the pandemic, several SARS-CoV-2 variants with mutations in RBD (and other) genes have emerged and spread globally, facilitating evasion of neutralizing antibody responses elicited by COVID-19 vaccination or previous infection. We have reported previously that plasma samples collected from individuals after primary SARS-CoV-2 infection show characteristic serological profiles, with preferential binding to the RBD of the infecting variant, and that the breadth of antibody binding to other variants improves over time with ongoing affinity maturation (10). Thus, SARS-CoV-2 variant serotyping may be used to determine exposure to SARS-CoV-2 variants in epidemiological studies.

SARS-CoV-2 infection during pregnancy has been associated with more severe COVID-19 disease (11), preterm delivery (12), increased fetal growth restriction (FGR), and lower birth weight in some populations (13), but not others (14, 15). Infection with Alpha or Delta variants may be associated with worse perinatal outcomes compared to infection with wildtype or Omicron variants (16-18). Intrauterine and direct neonatal SARS-CoV-2 infection is rare (19). There is a lack of data from resource limited settings in Africa related to effects of SARS-CoV-2 in pregnancy on birth outcomes and infant growth. Here, we use serological approaches to compare pre-pandemic serologic responses to SARS-CoV-2 and other coronaviruses in Ugandan and U.S. adults, and to retrospectively identify SARS-CoV-2 infections in a pregnancy cohort in eastern Uganda, describe serologic responses to SARS-CoV-2 variants, and correlate SARS-CoV-2 infection in women with reported symptoms and outcomes of infection in mothers and infants.

## Methods

### Parent studies

We leveraged data and samples from pregnant women and a subset of infants born to them enrolled in malaria chemoprevention trials that were ongoing during the SARS-CoV-2 pandemic (**Figure 1**). In the maternal trial, “Optimal chemopreventive regimens to prevent malaria and improve birth outcomes in Uganda” (NCT04336189), pregnant women are randomized to receive one of three monthly intermittent preventive treatment in pregnancy (IPTp) regimens: sulfadoxine-pyrimethamine (SP); dihydroartemisinin-piperaquine (DP); or DP+SP. Inclusion criteria are viable singleton pregnancy at 12-20 weeks gestational age, not living with human immunodeficiency virus (HIV), ≥16 years of age, resident of Busia District (in the Eastern Region of Uganda), and willingness to deliver in hospital. Exclusion criteria are long QT syndrome or active medical issue requiring inpatient treatment. At each clinic visit, a questionnaire was administered to collect information on presence or history of SARS-CoV-2 related symptoms (fever, fatigue/malaise, nausea, vomiting, diarrhea, cough, headache, rhinorrhea, abdominal pain, loss of taste/smell). Participants were referred for SARS-CoV-2 PCR or rapid antigen testing at the discretion of medical staff and subject to availability of COVID-19 testing materials. Socioeconomic data was collected for each mother at a home visit. Plasma samples are collected at enrollment and at delivery. Participants are tested for malaria parasites by PCR and blood smear at each monthly routine visit, and with blood smear at each unscheduled visit with fever. After delivery, placental tissue is evaluated by histopathology and for evidence of placental malaria, and birth outcomes are assessed. The maternal trial began enrolling in December 2020, and 429 participants had delivered by March 2022.

**Figure 1.**
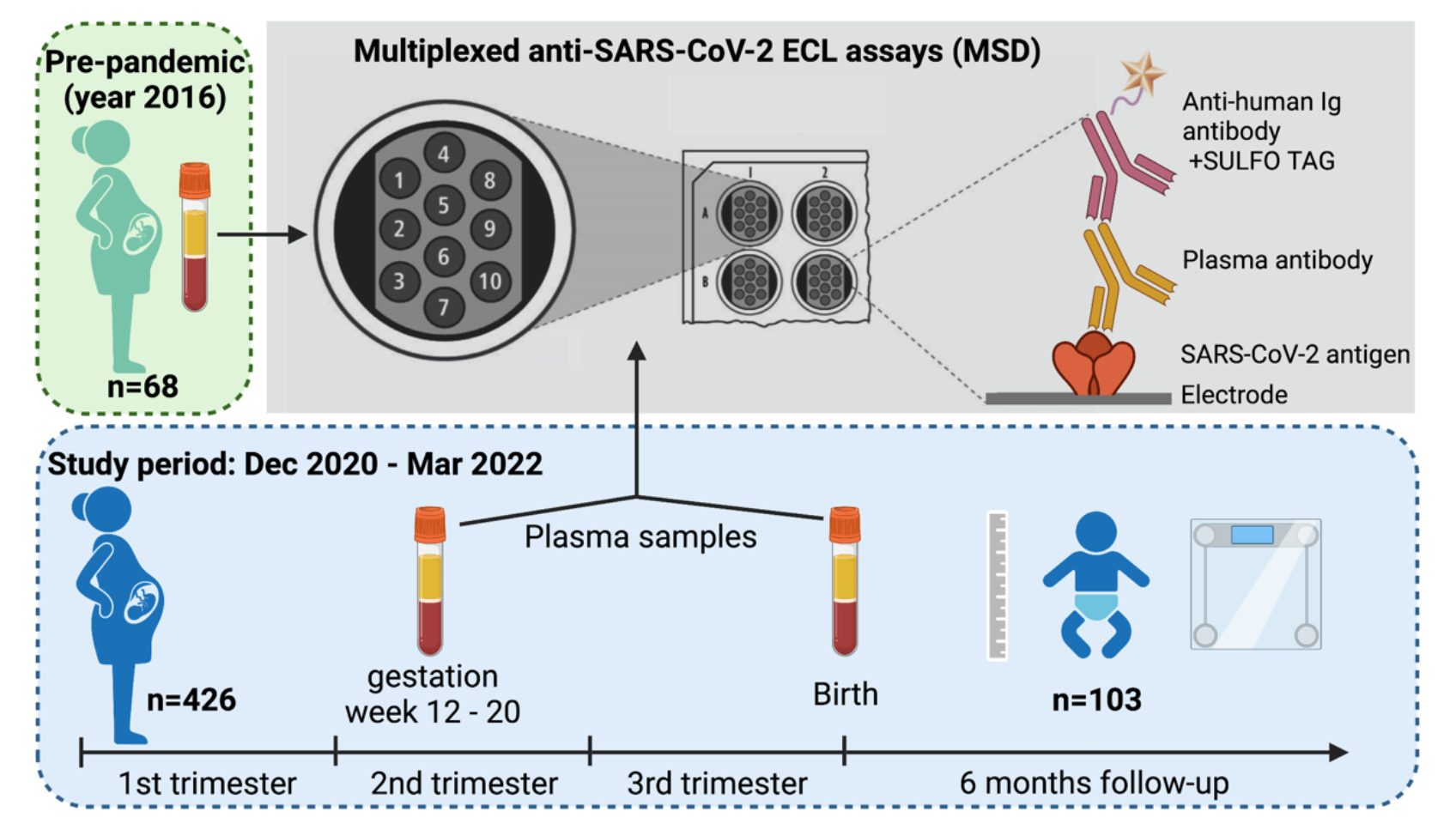
**Study Design.** We tested stored plasma from n=68 pregnant Ugandan women collected before the COVID-19 pandemic and n=426 (26 vaccinated against SARS-CoV-2 prior to or during study participation, 400 unvaccinated) pregnant Ugandan women enrolled in a malaria chemoprevention trial during the COVID-19 pandemic. Paired samples collected at enrollment (early 2^nd^ trimester) and at delivery were tested for SARS-CoV-2 IgG and IgM on the MSD platform to assess for SARS-CoV-2 infection in pregnancy and associations with clinical and birth outcomes. Of 400 unvaccinated women, SARS-CoV-2 infection status in pregnancy was able to be determined for n=320. Infant outcomes through 6 months of age were assessed in n=103 infants (84 with maternal SARS-CoV-2 status known) followed to 6 months of age.

Starting in October 2021, infants born to women enrolled in the maternal IPTp trial were enrolled in two separate studies. In the IMPACT (Infant malaria following IPTp with artemisinin-based combination therapy) study, infants born to the women in the IPTp trial are followed until one year of life. In an additional infant trial entitled “Enhancing immunity to malaria in young children with effective chemoprevention” (NCT04978272), infants born to the women in the IPTp trial are being randomized to receive IPT in childhood (IPTc) with monthly DP up to one year of age, monthly DP up to two years of age, or placebo. All children are followed up to four years of age. Inclusion criteria for both infant studies include infant’s mother enrolled in the maternal IPTp trial, resident of Busia district, and between 4 and 8 weeks of age at infant enrollment. Children are excluded if they have an active medical problem requiring frequent medical attention. Study infants receive all medical care at a dedicated study clinic at Masafu General Hospital. Participants are encouraged to come for unscheduled visits in the case of any febrile episode or other illness. Routine assessments are conducted every four weeks including collection of anthropometric data and collection of blood by finger prick for the detection of parasites by microscopy and quantitative PCR. N=134 infants born before March 2022 were enrolled in the infant studies.

### Sample Collection

From the maternal IPTp trial, paired plasma samples collected at enrollment (12-20 weeks of gestation) and at delivery were selected from a subset of 426 women with available samples from both time points who delivered before March 2022 (Figure 1). Stored plasma samples were also obtained from pre-pandemic clinical cohorts in California (91 samples collected in 2018 and 2019 from 45 adults ages 22-94 years enrolled in an influenza vaccination study) and Uganda (68 samples collected in 2016 from pregnant women enrolled in a previous malaria chemoprevention trial [NCT02793622]).

### SARS-CoV-2 Serology Testing

Plasma samples were tested for SARS-CoV-2-specific antibodies using the Meso Scale Discovery (MSD) electrochemiluminescence (ECL) platform. Plasma samples were heat-inactivated at 56 C for 30 minutes and tested in a 96-well plate format with MSD V-PLEX serology panels and instrumentation according to the manufacturer’s instructions. V-PLEX COVID-19 Coronavirus Panel 3 kits were used to detect IgG antibodies to SARS-CoV-2 N, RBD, and Spike antigens and to Spike proteins of SARS-CoV-1, MERS-CoV, and other endemic HCoVs including HCoV-OC43, HCoV-HKU1, HCoV-NL63, and HCoV-229E. V-PLEX SARS-CoV-2 Panel 8, 23, and 26 kits were used to determine IgG antibody concentrations of different SARS-CoV-2 variant Spike and RBD antigens, including Wuhan-Hu-1, Alpha, Beta, Gamma, Delta, Eta, and Omicron. We also used V-PLEX SARS-CoV-2 Panel 8 kits to determine IgM antibody concentrations to SARS-CoV-2 N, RBD, and Spike antigens. Plasma samples were analyzed at a 1:5,000 dilution in MSD diluent. Coronavirus-specific antibodies were detected with anti-human IgG or IgM antibodies conjugated to SULFO-TAG ECL labels and read with a MESO QuickPlex SQ 120 instrument. Cutoff values for positive antibody test results were determined for each antigen by testing the sera from Ugandan pre-pandemic control pregnant women and were defined as the mean plus five standard deviations of the results from the pre-pandemic specimens. IgG antibody binding ratios for Wuhan-Hu-1 RBD and viral variant RBDs were calculated and plotted only for specimens that were above the highest RBD cutoff value for positive to avoid distortion of ratios by samples without specific binding. Each MSD plate contained duplicates of a 7-point calibration curve with serial dilution of a reference standard and a blank well. Calibration curves were used to calculate antibody unit (AU) concentrations by backfitting ECL signals measured for each sample to the curve.

To test the consistency of serological results for Wuhan Spike using two different assays, a subset of samples were additionally tested using a Luminex multiplex bead assay assessing total IgG responses to the spike protein, as previously described (8).

### Classification of SARS-CoV-2 exposure status

Samples were defined as positive for SARS-CoV-2 IgM if the AU concentration of Spike or RBD IgM antibodies was above the positive threshold as defined above and positive for SARS-CoV-2 IgG if the AU concentration of Wuhan-Hu-1-specific Spike or RBD IgG, or any other variant Spike IgG, were above the positive threshold. Participants who were negative for IgG and IgM at both enrollment and delivery time points were defined as uninfected with SARS-CoV-2. Participants who were negative for both IgG and IgM at enrollment but positive for either or both IgG and IgM at delivery were defined as seroconverted and assumed to be infected during their study participation (i.e., between the second or third trimester of pregnancy). Participants with positive IgM (with or without positive IgG) at enrollment (between weeks 12 and 20 of gestation), were classified as likely infected with SARS-CoV-2 during early pregnancy, given the expected decay of IgM within 60 days after infection (10). If a participant was IgG positive at enrollment and Spike IgG AU increased ≥4-fold between enrollment and delivery, they were considered to have a “boosted” response and likely re-infected during the study period (20). Participants who seroconverted, boosted, or had early pregnancy exposure were classified as infected during pregnancy. Presumed variant of exposure was defined as the variant with the lowest Wuhan-Hu- 1:variant RBD IgG AU ratio, with ratio <1, indicating a stronger response to the variant than to the early Wuhan-Hu-1 strain or other variants.

### Statistical Analysis

All analyses were performed using the R statistical software, version 4.2.2 (https://www.r-project.org/). Box-whisker plots of serological data show median (horizontal line), interquartile range (box), and the whisker ends as the most extreme values within 1.5 times the interquartile range. Differences in antibody concentrations to coronavirus antigens between participant cohorts were tested using a two-sided Wilcoxon rank sum test and p-values <0.05 were considered statistically significant. In IgG Wuhan-Hu-1 to variant RBD ratio plots, a regression line was added using ggplot2 to show the general trend of IgG binding to variants over time. Participant characteristics and clinical outcomes were compared between participants who were and were not infected with SARS-CoV-2 during pregnancy using Chi-square tests for categorical variables, Student’s t-tests for continuous variables if normally distributed, and Mann-Whitney U tests or Kruskal Wallis tests for continuous variables with non-normal distributions. To evaluate factors associated with infant stunting at multiple time points during the first 24 weeks of life, we fit linear mixed effects regression models with random intercepts for each infant for the continuous outcomes of length-and weight-for-age Z score (per WHO growth standards (21)), with SARS-CoV-2 during early or late pregnancy as the primary exposure of interest, adjusting for maternal age, education, gravidity, body mass index (BMI), infant sex, infant age, placental malaria, and infant length at birth using the ’lmer’ function of the package lme4 (22). Models were also fit with interaction terms between SARS-CoV-2 infection and infant age, and between SARS-COV-2 infection and placental malaria.

### Study Approval

Recruitment of patients, documentation of written informed consent prior to participation, collection of blood specimens, and experimental measurements were carried out with Institutional Review Board approval at Stanford University (Stanford, CA, USA), UCSF (San Francisco, CA, USA) and Infectious Diseases Research Collaboration (Kampala, Uganda).

### Data Availability

Underlying data and supporting analytic code for the article can be accessed upon request.

## Results

### Ugandan women did not show elevated pre-pandemic concentrations of IgG to Spike proteins of SARS-CoV-2 and other coronaviruses

One hypothesis to explain the relatively mild clinical impact of the COVID-19 pandemic in Africa is that, compared to other regions, higher levels of endemic coronavirus population exposure may have led to increased pre-existing cross-protective immunity to SARS-CoV-2 in Africa. We therefore compared concentrations of antibodies to SARS-CoV-2 antigens and Spike proteins of other human coronaviruses in pre-pandemic samples from Ugandan and Californian study participants (Figure 2). IgG concentrations to SARS-CoV-2 RBD and Spike as well as to the Spike protein of all other tested human coronaviruses, including SARS-CoV-1, MERS, HCoV-229E, HCoV-HKU1, HCoV-NL63, and HCoV-OC43, were either not significantly different between Ugandan and Californian study cohorts or significantly higher in the Californian group (Figure 2A). In contrast, IgG concentrations to SARS-CoV-2 N and IgM antibody concentrations to SARS-CoV-2 N, RBD, and Spike antigens were significantly higher in the Ugandan study cohort (Figure 2B).

**Figure 2.**
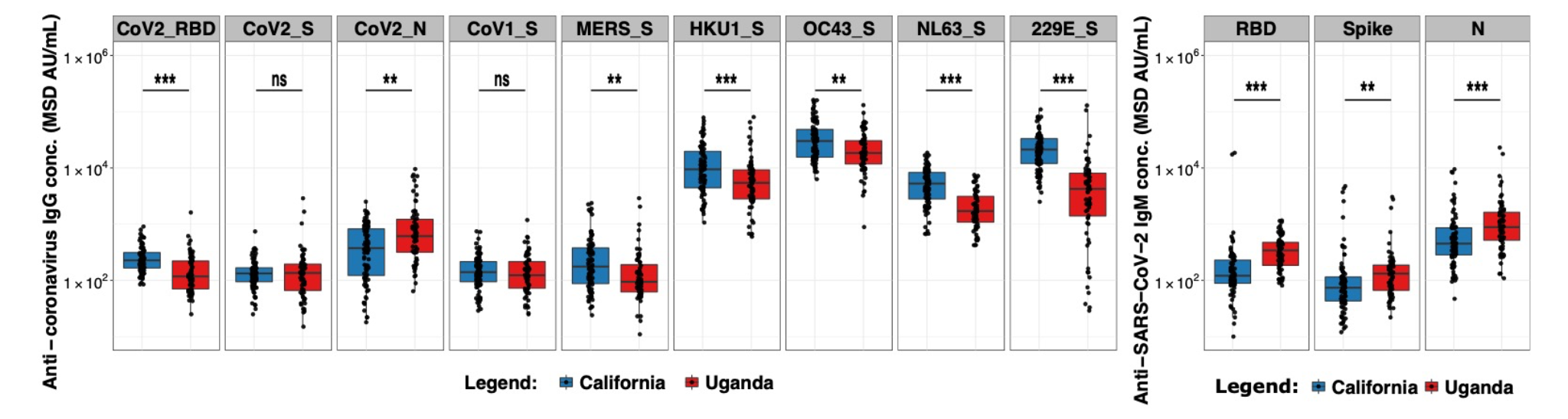
**Low levels of anti-SARS-CoV-2 IgG concentrations in pre-pandemic Ugandan study cohort.** Anti-coronavirus IgG (A) and IgM (B) antibody concentrations in MSD AU/mL are shown for pre-pandemic samples from a Californian (blue, n=91) and a Ugandan (red, n=68) study cohort. Box-whisker plots show the interquartile range as the box and the whisker ends as the most extreme values within 1.5 times the interquartile range below the 25% quantile and above the 75% quantile. Wilcoxon rank sum test was used to test for significant differences, with *p<0.05, **p<0.01, ***p<0.001.

### High levels of SARS-CoV-2 infection were detected between late 2020 and early 2022

Underreporting of SARS-CoV-2 infection and COVID-19 related death has been suspected in African countries. Previous serological investigations have indicated high levels of exposure of African populations to SARS-CoV-2 (4). We tested paired plasma samples from enrollment and delivery from 426 pregnant Ugandan women for the presence of anti-SARS-CoV-2 N, RBD, and Spike IgG (Figure 3A) and IgM (Figure 3B) antibodies using multiplexed MSD ECL assays. There was high correlation between anti-SARS-CoV-2 Spike IgG as measured by the MSD and Luminex assays (Figure S1). Of the 426 study participants, 26 reported being vaccinated and were excluded from further SARS-CoV-2 exposure analysis. Of 400 unvaccinated participants, 220 were negative for anti-Spike and anti-RBD IgG and IgM at enrollment. Of these, 128 remained antibody negative at delivery and were presumed to have not been infected with SARS-CoV-2 in pregnancy, and 92 were anti-Spike and/or anti-RBD antibody positive at delivery and presumed to have experienced SARS-CoV-2 infection during pregnancy. Of the 180 who were positive for IgG and/or IgM at enrollment, 128 (71.1%) were positive for IgG only, 12 (6.7%) for IgM only, and 40 (22.2%) for both IgG and IgM. The 52 who were positive for IgM only or IgM and IgG were presumed to have been infected with SARS-CoV-2 early in pregnancy prior to study enrollment. Of the 128 who were positive for only IgG at enrollment, 48 had antibodies boosted by a factor of 4 or greater at delivery and were presumed to have been re-infected during late pregnancy. Timing of infection could not be determined in the 80 with positive IgG and negative IgM at enrollment and no boosting at time of delivery, since infection may have occurred prior to pregnancy or early in pregnancy, and these participants were excluded from birth and infant outcome analysis. Ultimately, we presumed that 128 participants remained uninfected with SARS-CoV-2, and 192 were infected with SARS-CoV-2 at some point during their pregnancy (Figure 4).

**Figure 3.**
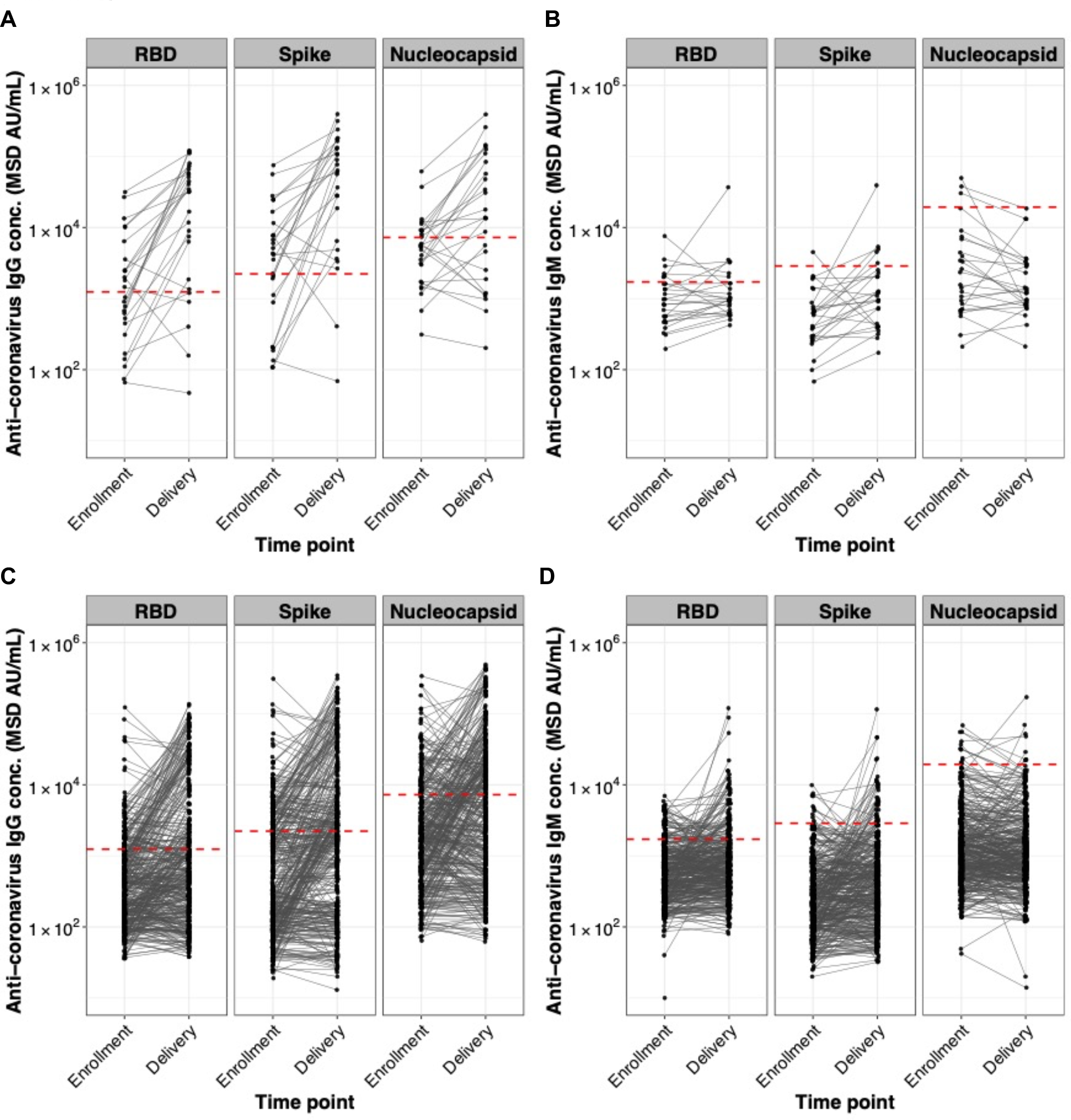
**High levels of SARS-CoV-2 infection in Ugandan pregnancy cohort during COVID-19 pandemic.** Anti-SARS-CoV-2 RBD, Spike, and N IgG (A,C) and IgM (B,D) antibody responses are shown for samples taken at study enrollment and delivery from pregnant women in Uganda who were vaccinated (A,B) and unvaccinated (C, D) against SARS-CoV-2 prior to or during study participation. Paired samples are connected with black lines. Dashed red lines indicate cutoff values for positive as determined by testing pre-pandemic samples from Ugandan women collected in 2016. Pre-pandemic background levels for anti-SARS-CoV-2 IgM antibodies were high compared to pandemic responses, particularly for anti-N.

**Figure 4.**
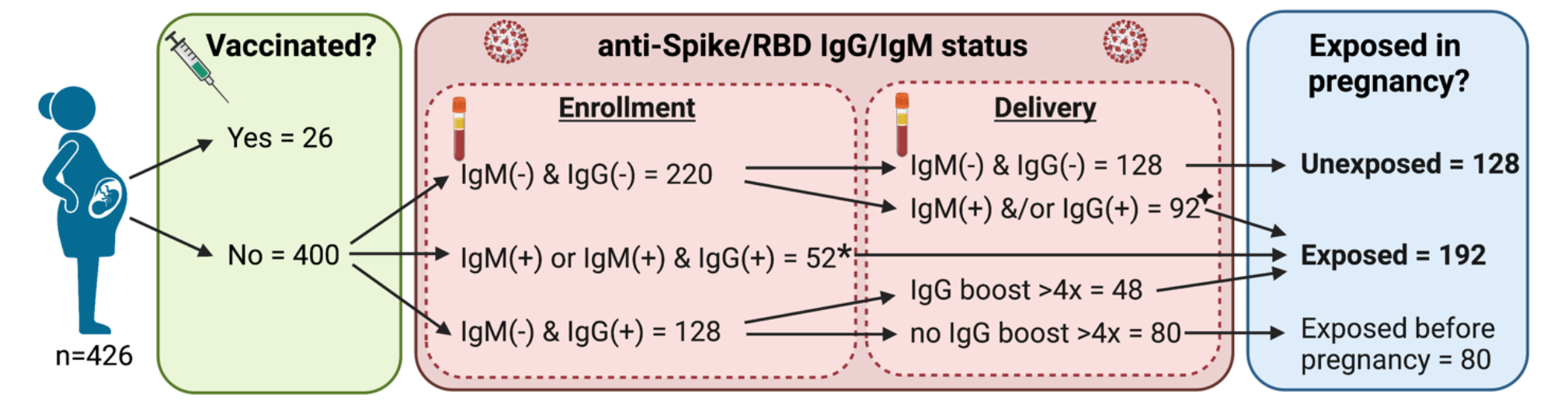
**SARS-CoV-2 Exposure During Pregnancy.** N=426 participants enrolled in a trial of malaria chemoprophylaxis in pregnancy in Uganda between December 2020 and February 2022 had stored plasma samples collected at enrollment (between 12 and 20 weeks gestation) and at time of delivery tested for SARS-CoV-2 Spike antigen and receptor binding domain (RBD) IgG and IgM using the MesoScale Discovery (MSD) platform. Of n=400 participants who remained unvaccinated to SARS-CoV-2 throughout study participation, 220 were negative for IgG and IgM to both Spike and RBD at enrollment. Of these, 128 remained negative at delivery and were presumed to have not been exposed to SARS-CoV-2 in pregnancy, and 92 were positive at delivery and were classified as seroconverted indicating SARS-CoV-2 infection during late^✦^ pregnancy. Of the 180 who were positive for IgG and/or IgM at enrollment, 128 were positive for IgG only, 12 for IgM only, and 40 for both IgG and IgM. The 52 who were positive for IgM only or IgM and IgG were presumed to have been infected with SARS-CoV-2 early* in pregnancy prior to study enrollment. Of the 128 who were positive for only IgG at enrollment, 48 had IgG antibodies boosted by a factor of 4 or greater at delivery and were presumed to have been re-infected during study participation. N=80 had only IgG positive at enrollment with negative IgM and no boosting at time of delivery, thus infection occurred prior to pregnancy or shortly thereafter, and timing is unable to be determined. Therefore, these participants were excluded from birth and infant outcome analysis.

### The breadth of SARS-CoV-2 variant RBD IgG binding improved over time

We showed previously that following primary SARS-CoV-2 infection, plasma IgG antibodies preferentially bind to the RBD of the infecting variant (10). Here, we determined plasma IgG concentrations binding to the RBDs of Wuhan-Hu-1; five SARS-CoV-2 variants including Alpha, Beta, Gamma, Delta, and Eta; and three Omicron subvariants (BA.1, BA.1.1, and BA.2) using multiplexed MSD ECL assays (Figure 5A). To evaluate differences in variant RBD binding over time, we compared the ratios of anti-RBD IgG concentrations for Wuhan-Hu-1 and other variants (Figure 5B). Based on the serological profile of RBD variant binding we identified putative Alpha, Delta, and Eta variant exposure mainly in early to mid-2021. Interestingly, IgG binding ratios, which showed a clear variant preference in the enrollment period, changed over time toward equal binding to Wuhan-Hu-1 and variants at delivery, indicating an increase in the breadth of the antibody response. Probable exposure to different Omicron subvariants in late 2021 and early 2022 was notable in that it was not accompanied by the expected preference of Omicron over Wuhan-Hu-1 RBD binding.

**Figure 5.**
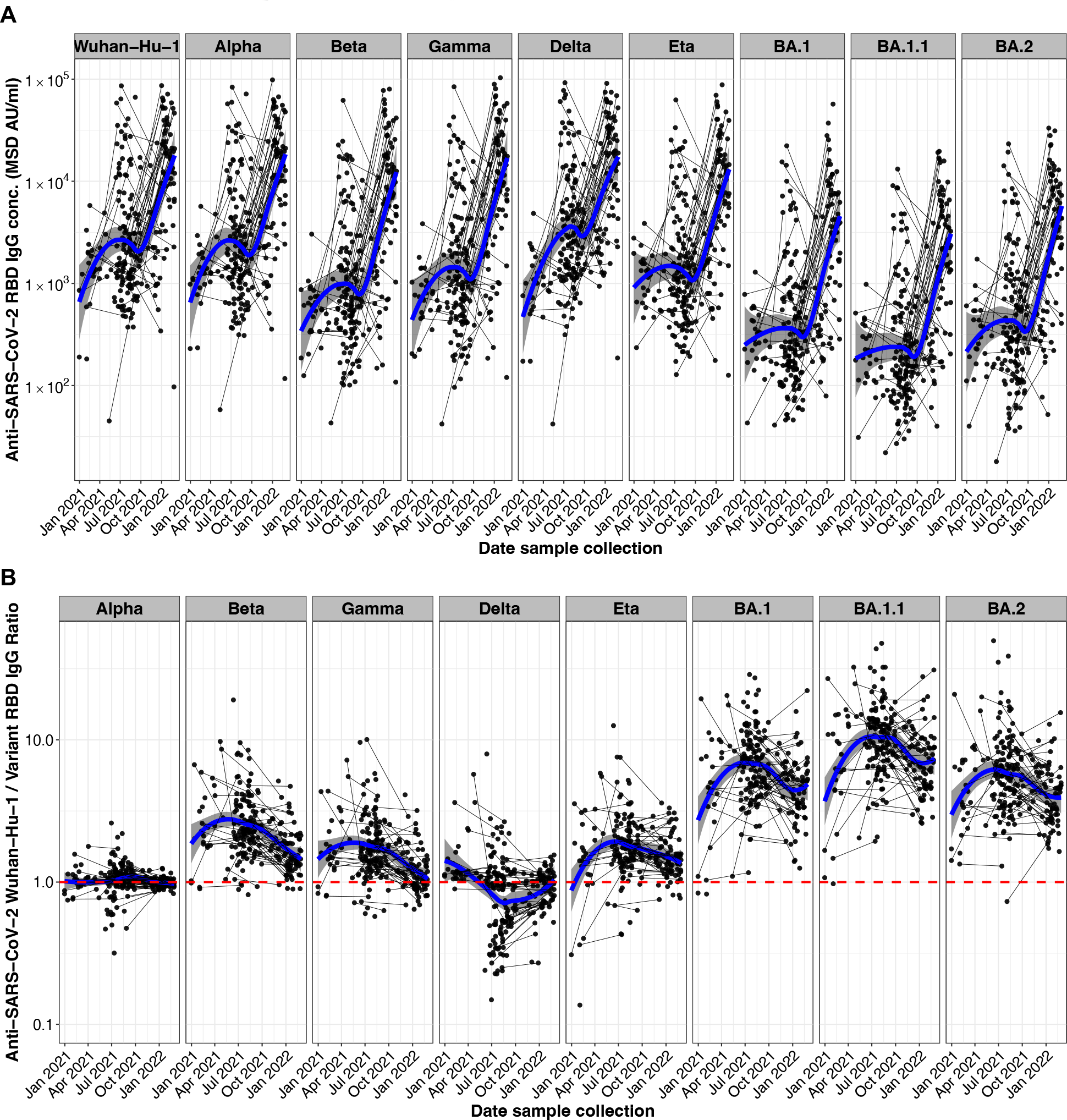
**Exposure of Ugandan pregnancy cohort to different SARS-CoV-2 variants.** Anti-SARS-CoV-2 Wuhan-Hu-1 and viral variant RBD IgG responses in samples from pregnant women in Uganda between late 2020 and early 2022 are shown. Paired samples from study participants at enrollment and delivery timepoints are connected with a black line. The blue locally estimated scatterplot smoothing (LOESS) regression line shows the trend of the development of the antibody response over time. A) Anti-RBD IgG concentrations in MSD AU/mL. B) Ratios of anti-Wuhan-Hu-1 to variant RBD IgG concentration. The red dashed line indicated a ratio of 1 in which the IgG variant response is equal to the anti-Wuhan-Hu- 1 response; below the red line indicates preferential binding to the variant RBD, whereas above the red line indicates preferential binding to Wuhan-Hu-1 RBD.

### Exposure to SARS-CoV-2 variants and reported symptoms in the pregnancy cohort

Increased numbers of SARS-CoV-2 infections were reported in the general Ugandan population during Alpha, Delta, and Omicron waves (Figure 6A), and was temporally correlating with SARS-CoV-2 variant-specific serological ratios in our study cohort (Figure 5B). Reports of upper respiratory infections (URIs) and cough increased during the Delta wave in June-July 2021 and Omicron wave in December 2021-January 2022. Report of fever did not similarly increase during these waves, however URI, fever, and cough all were increased during a period of high malaria incidence in February 2021 (Figure 6B). Cumulative numbers of SARS-CoV-2 infections were reflected in the increased seropositivity rate from less than 10% at enrollment in December 2020 to 100% in January 2022 (Figure 6C).

**Figure 6.**
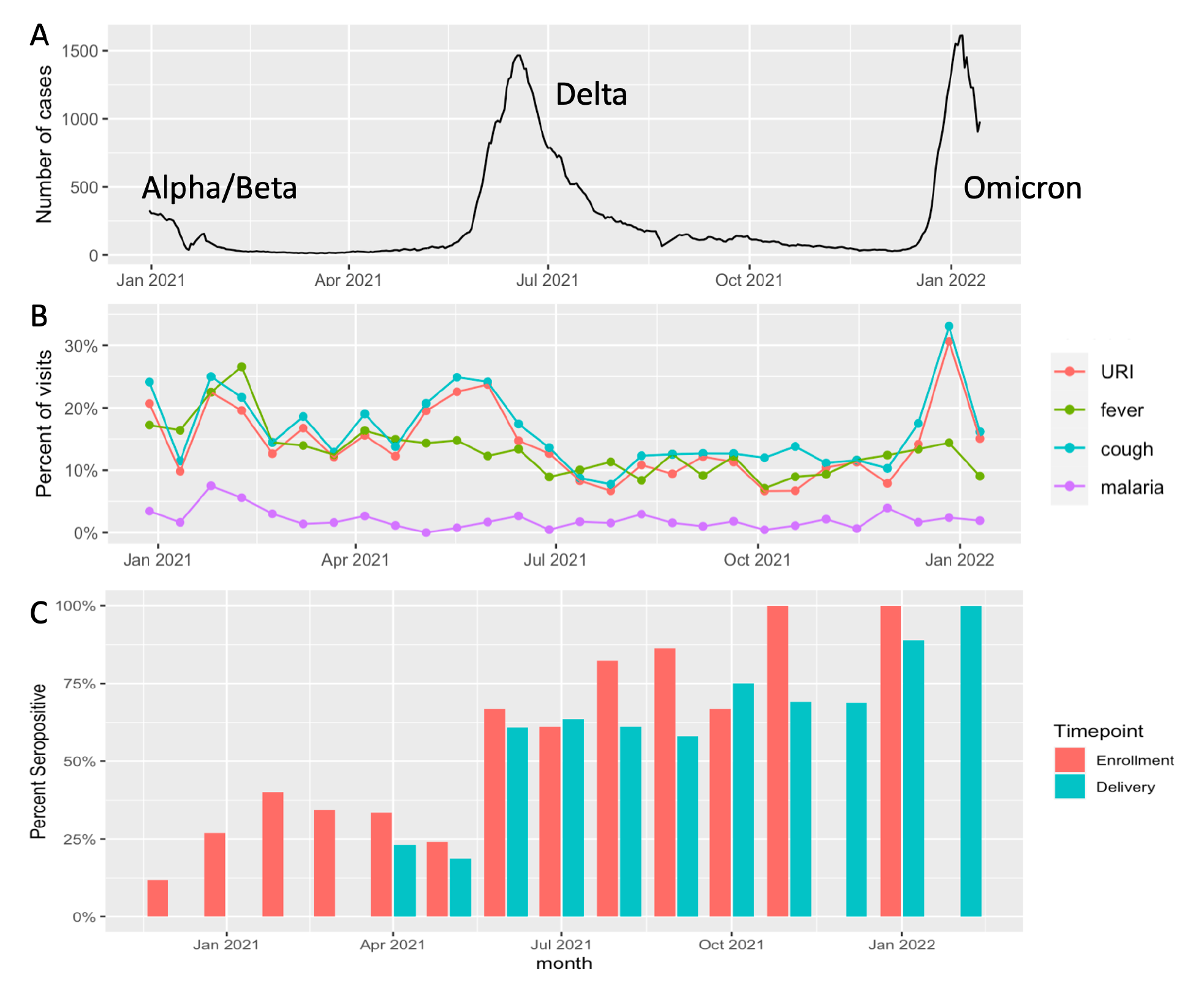
**(A-C). SARS-CoV-2 surges in Uganda (A) associated with symptoms (B) and seropositivity at enrollment (C) over time.** A. 7-day rolling average of daily new COVID-19 cases reported in Uganda (obtained from Our World in Data database, https://ourworldindata.org/coronavirus/country/uganda). One week in late August 2021 when >21,000 cases were reported in a single day is omitted. Surges are labeled with associated SARS-CoV-2 variants. B. Percent of study visits in each 2-week period from December 2020-February 2022 in which participants reported upper respiratory tract infection (URI), fever, cough, or were diagnosed with malaria (fever+positive blood smear with Plasmodium falciparum). Numbers of visits with reported cough or upper respiratory infections (URI) increased during the Delta and Omicron waves in Uganda. C. The proportion of seropositive tests at either enrollment (red) or delivery (blue) visits in each month increased from 12% in December 2020 to 100% in February 2022.

Given the high rate of reported cough during the study period, we examined boosting of antibodies to human coronaviruses other than SARS-CoV-2 (Figure 7). As described above, many participants experienced rises in antibody levels against SARS-CoV-2 N, RBD, and Spike antigen throughout the course of the study period; many participants also showed increases in antibody levels to SARS-CoV-1 and MERS, likely due to antibody cross-reactivity with SARS-CoV-2 antigens. A smaller subset of participants showed evidence of at least 4-fold boosting of antibodies to the Spike protein of seasonal HCoVs NL63, HKU1, OC43 and 229E (n=8, 5, 11, and 29, respectively), suggesting that these coronaviruses may have also contributed to cough in some members of the cohort.

**Figure 7.**
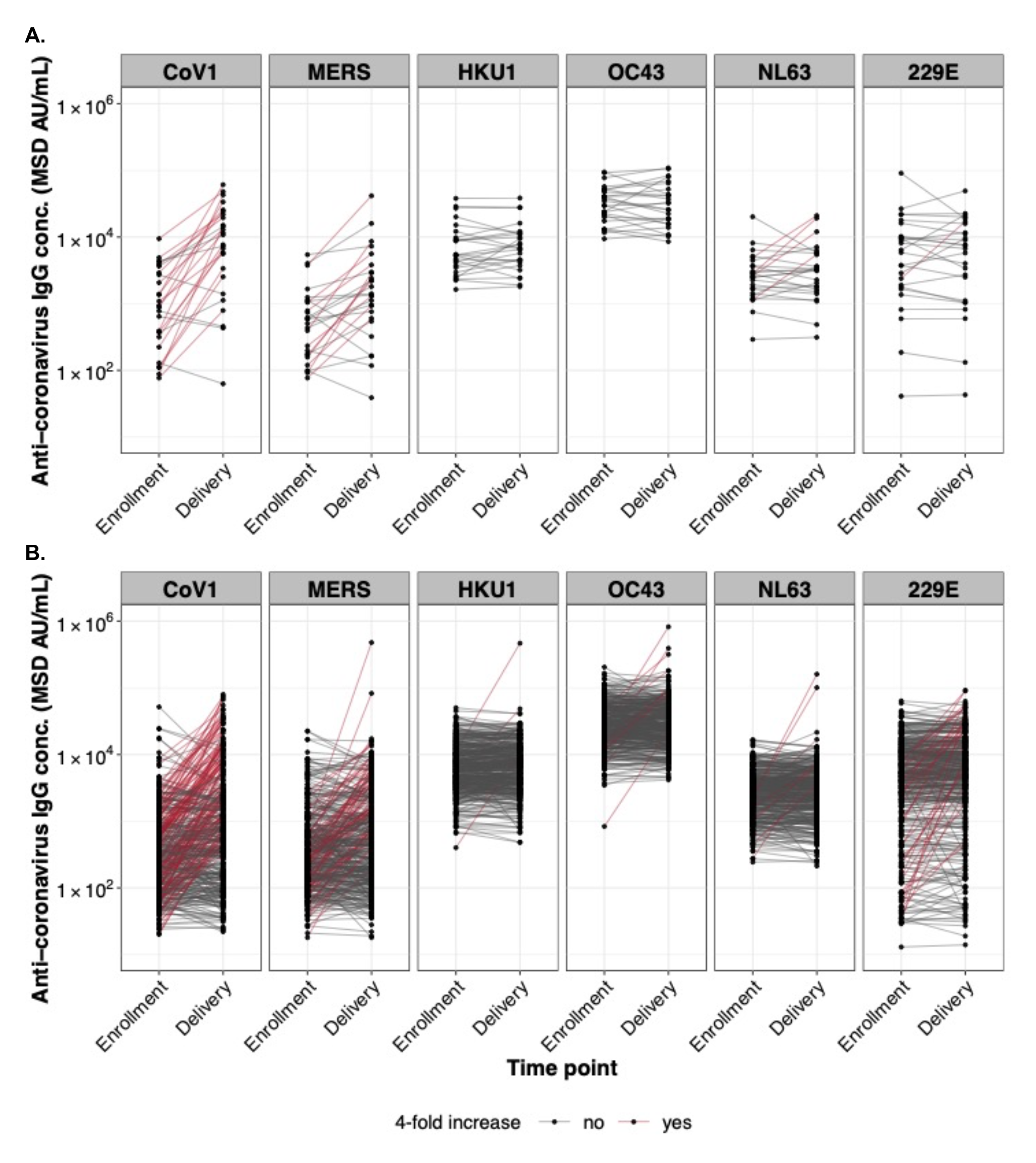
**Antibody cross-reactivity in samples from Ugandan pregnancy cohort to different coronavirus Spike proteins.** Antibody responses to the Spike protein of SARS-CoV-1, MERS-CoV, and other endemic HCoVs including HCoV-OC43, HCoV-HKU1, HCoV-NL63, and HCoV-229E are shown for samples taken at study enrollment and delivery from vaccinated (A) and unvaccinated (B) pregnant women in Uganda. Lines connecting paired samples are red for participants who exhibited boosting of IgG levels by a factor of 4 or greater between enrollment and delivery.

### Baseline characteristics and clinical manifestations associated with SARS-CoV-2 infection in pregnancy

We next compared characteristics of pregnant women who were and were not infected with SARS-CoV-2 during pregnancy (Table 1). There was no statistically significant difference in age, gravidity, maternal BMI at enrollment, education, or malaria incidence per person year during pregnancy. There was a trend towards lower malaria parasitemia, as measured by qPCR at enrollment, in SARS-CoV-2-infected participants compared to uninfected participants (Table 1; 54.7% vs 64.8%; p=0.07). However, a higher proportion of participants living in modern houses were found to be infected with SARS-CoV-2 (Table 1; OR 1.72, 95% CI 1.05-2.85, p=0.03) compared to those living in traditional houses. On the other hand, a lower proportion of participants living in modern houses had malaria parasitemia at enrollment (OR 0.44, 95% CI 0.29-0.67, p<0.001). These findings suggest that the environmental factors associated with malaria exposure are different from those associated with SARS-CoV-2 exposure.

**Table 1.**
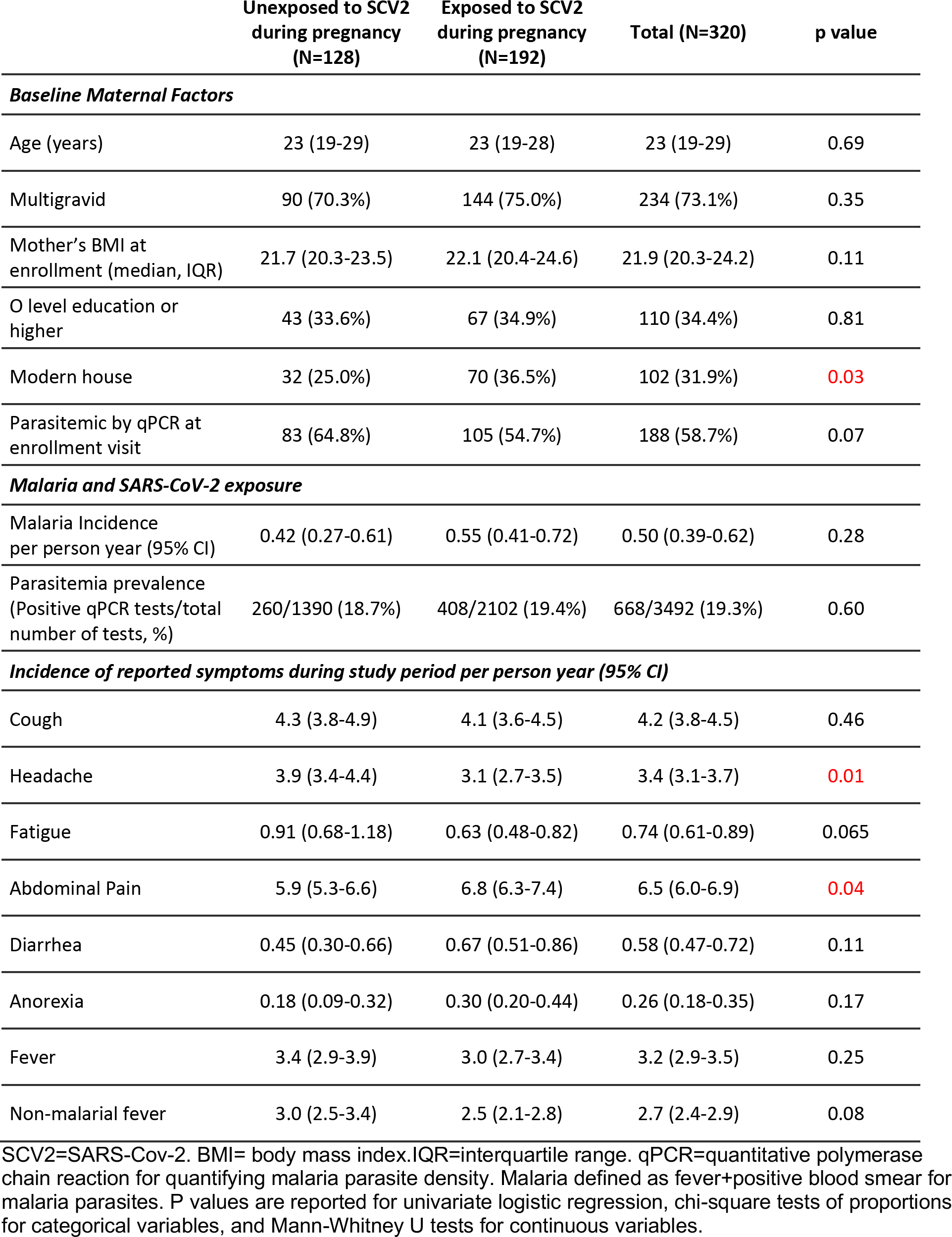
Maternal factors and symptoms associated with SARS-CoV-2 exposure in pregnancy.

At the group level, increases in cough and URI corresponded to COVID-19 waves. However, individual symptom categories such as cough and fever were not associated with seroconversion, consistent with findings in non-pregnant adults in the same setting (8). Abdominal pain was more prevalent in SARS-Co-2 infected participants, but headache was more common in SARS-Co-2 uninfected participants (Table 1). Despite the high rates of seroconversion for anti-SARS-CoV-2 antibodies during the study period, only three participants were tested by rapid antigen test or PCR during the study period, and only one participant was diagnosed with COVID-19 by rapid antigen test. This participant presented in the third trimester to study clinic with subjective fever, myalgias, and orolabial sores, but no cough or dyspnea. Though she was afebrile and hemodynamically stable with oxygen saturation 97%, she was hospitalized for 3 days for monitoring out of an abundance of caution. She was treated with azithromycin, paracetamol, vitamin C and zinc. All symptoms resolved and she went on to deliver a healthy, normal weight infant at 40 weeks of gestation. No participants had severe respiratory illness during study participation. There was one death in a participant who seroconverted with presumed Delta infection, but this death was due to a surgical delivery complication unrelated to COVID-19. One participant with likely first trimester Alpha variant COVID-19 experienced transverse myelitis with unclear etiology; she was treated with steroids and recovered.

### SARS-CoV-2 infection during pregnancy was associated with stunting and lower length-for-age Z scores in early infancy

We next sought to determine whether SARS-CoV-2 infection during pregnancy was associated with adverse birth and infant outcomes, as infections during pregnancy have been associated with adverse birth outcome in some studies (12, 13). Though the proportion of spontaneous abortion, stillbirth, and preterm birth in infants born to infected mothers were marginally higher than uninfected mothers, these differences did not reach statistical significance (Table 2). There was no difference in median newborn weight or length, and though low birth weight, C-section delivery, placental malaria and intervillous inflammation on histology were marginally higher in infants born to SARS-CoV-2-uninfected mothers compared with infected mothers, again these differences did not reach statistical significance.

**Table 2.** Delivery outcomes associated with SARS-CoV-2 exposure in pregnancy.

| **Delivery outcomes** | **Not Exposed in**<br>**Pregnancy (N=128)** | **Exposed in**<br>**Pregnancy (N=192)** | **Total (N=307)** | **p value** |
| --- | --- | --- | --- | --- |
| **Spontaneous abortion, n (%)** | 2 (1.6%) | 4 (2.1%) | 6 (1.9%) | 0.743 |
| **Stillbirth, n (%)** | 3 (2.4%) | 6 (3.2%) | 9 (2.9%) | 0.682 |
| **Preterm birth (<37 weeks), n (%)** | 5 (4.1%) | 10 (5.5%) | 15 (4.9%) | 0.582 |
| **Gestational age at birth (weeks),**<br>**median (IQR)** | 39.0 (38.0-40.0) | 39.0 (38.0-40.0) | 39.0 (38.0-40.0) | 0.475 |
| **Newborn weight (g), median (IQR)** | 3000 (2800, 3285) | 3000 (2758, 3300) | 3000 (2770, 3300) | 0.944 |
| **Newborn length (cm), median (IQR)** | 47.0 (45.0, 48.0) | 47.0 (45.0, 48.0) | 47.0 (45.0, 48.0) | 0.646 |
| **Low birth weight (<2500 g), n (%)** | 11 (9.1%) | 12 (6.6%) | 23 (7.6%) | 0.421 |
| **Small for gestational age (<10<sup>th</sup>**<br>**percentile), n (%)** | 21 (17.4%) | 30 (16.5%) | 51 (16.8%) | 0.843 |
| **C-section delivery, n (%)** | 20 (16.0%) | 35 (18.6%) | 55 (17.6%) | 0.551 |
| **Male infant, n (%)** | 54 (43.2%) | 96 (50.8%) | 150 (47.8%) | 0.187 |
| **Mother's WBC at delivery (cells/μl),**<br>**median (IQR)** | 10470 (7117, 12865) | 10820 (8710, 13395) | 10650 (8260, 13230) | 0.106 |
| **Placental Malaria, n (%)** | 36 (30.0%) | 45 (26.9%) | 81 (28.2%) | 0.571 |
| **Intervillous inflammation present, n**<br>**(%)** | 21 (17.5%) | 21 (12.6%) | 42 (14.6%) | 0.244 |

WBC=white blood count. Placental malaria defined as malaria parasites or pigment on histology. P values are reported for chi-square tests of proportions for categorical variables, and Mann-Whitney U tests for continuous variables.

However, among the 84 infants who were enrolled in the infant study and whose mothers’ SARS-CoV-2 seroconversion status in pregnancy was known, statistically significant associations were observed between SARS-CoV-2 exposure during pregnancy and infant length-for-age Z scores and stunting at 12 weeks of age (Table 3). Birth length and length-for-age Z scores were lower in infants born to mothers infected with SARS-CoV-2 in early pregnancy (positive IgM at enrollment) compared to infants born to uninfected mothers or mothers infected with SARS-CoV-2 late in pregnancy (Figure 8).In linear mixed effects models with random intercepts for each participant, SARS-CoV-2 infection in early pregnancy was associated with lower infant length-for-age Z scores (Coef.=-1.07, 95%CI −1.79 – −0.35, p=0.003) in the first six months of life when adjusting for maternal and infant factors (Table 4). Lower maternal gravidity was associated with increased infant length-for-age Z scores (Coef.= −0.24, 95%CI −0.46- −0.02, p=0.030), and higher maternal BMI and increased infant birth length was associated with increased length-for-age Z scores (Coef.= 0.07, 95% CI 0.00-0.14, p=0.047; Coef.=0.15, 95%CI 0.04-0.26, p=0.009). Placental malaria (Coef.=-0.73, 95%CI −1.27- −0.18, p=0.009) and increasing infant age (Coef.=-0.01, 95%CI −0.01- −0.00, p=0.001) were associated with lower weight-for-age Z scores, while higher maternal BMI and increased infant birth length were associated with increased weight-for-age Z scores (Coef.= 0.13, 95% CI 0.07-0.19, p<0.001; Coef.=0.12, 95%CI 0.02-0.22, p=0.020).

**Figure 8.**
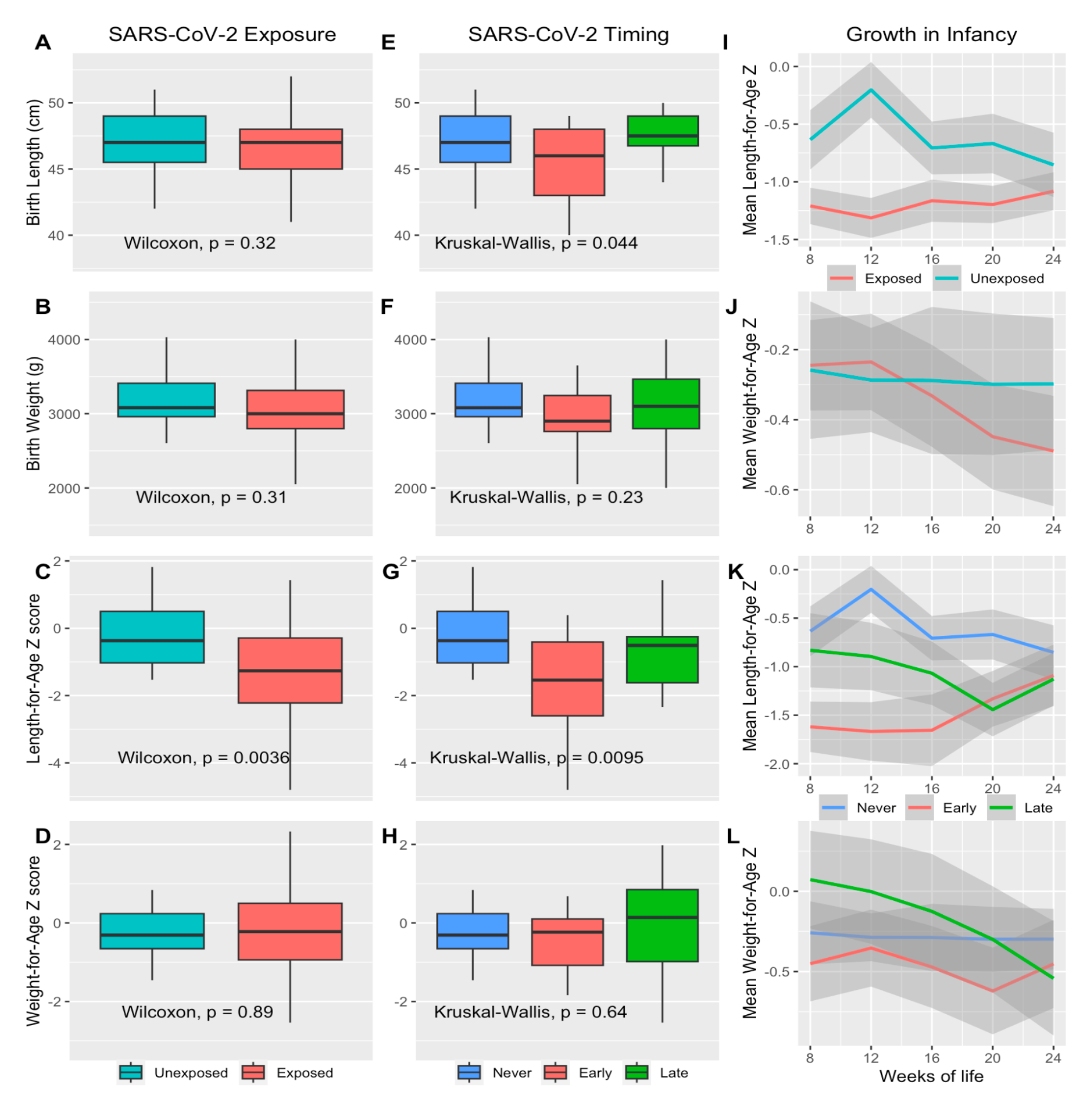
**Gestational SARS-CoV-2 exposure and infant growth.** Length- and Weight-for-age Z scores determined from WHO reference standards. Boxplots revealing infant length at birth, infant weight at birth, length-for-age Z score at age 12 weeks, and length-for-age Z score at age 12 weeks comparing SARS-CoV-2 exposed and unexposed (A-D) and timing of exposure (E-H). Panels I-L show mean and 95% CI (shaded region) of growth Z scores from 8 to 24 weeks of life comparing SARS-CoV-2 exposure (I-J) and timing of exposure (K-L). Exposed=exposed to SARS-CoV-2 during pregnancy as determined by seroconversion, positive Spike and/or RBD IgM at enrollment indicating recent infection, and/or boosting of IgG levels by a factor of 4 or greater between enrollment and delivery. Early infection=IgM positive at enrollment indicating recent infection in first trimester. Late infection=IgM and IgG negative at enrollment and IgM and/or IgG positive at delivery. Unexposed or never infected=remaining IgG and IgM negative at both enrollment and delivery.

**Table 3.** Infant growth outcomes associated with SARS-CoV-2 exposure during pregnancy.

|  | Not Exposed in Pregnancy<br>(N=19) | Exposed in Pregnancy<br>(N=65) | Total (N=84) | p value |
| --- | --- | --- | --- | --- |
| Height in cm at 12 weeks of age,<br>median (IQR) | 59.6 (58.0, 61.5) | 58.0 (55.0,60.0) | 58.0 (55.0,60.0) | 0.008 |
| Weight in kg at 12 weeks of age,<br>median (IQR) | 5.9 (5.3, 6.2) | 5.7 (5.3, 6.3) | 5.7 (5.3, 6.3) | 0.932 |
| Stunting at 12 weeks of age, n (%) | 0 (0.0%) | 21 (32.3%) | 21 (25.0%) | 0.002 |
| Underweight at 12 weeks of age, n<br>(%) | 0 (0.0%) | 3 (4.6%) | 3 (3.6%) | 1.000 |

IQR=interquartile range. Stunting=length-for-age Z score < -2 based on WHO Child Growth Standards. Underweight=weight-for-age Z score<-2 based on WHO Child Growth Standards. P values are reported for Fisher exact tests of proportions for categorical variables, and Mann-Whitney U tests for continuous variables.

**Table 4.** Linear mixed effects regression models to identify the effects of SARS-CoV-2 in pregnancy and other maternal and infant factors on length-and weight-for-age Z scores through 24 weeks of life

| *Predictors* | **Length-for-age Z Score** |  |  |  | **Weight-for-age Z Score** |  |  |  |
| --- | --- | --- | --- | --- | --- | --- | --- | --- |
|  | *Est* | *std. Error* | *CI* | *p* | *Est* | *std. Error* | *CI* | *p* |
| SARS-CoV-2<br>(Ref=Uninfected) |  |  |  |  |  |  |  |  |
| Infection in early pregnancy | -1.07 | 0.37 | -1.79 – -0.35 | **0.003** | -0.48 | 0.33 | -1.12 – 0.16 | 0.142 |
| Infection in late pregnancy | -0.30 | 0.36 | -1.00 – 0.41 | 0.409 | 0.20 | 0.32 | -0.42 – 0.83 | 0.526 |
| Gravidity | -0.24 | 0.11 | -0.46 – -0.02 | **0.030** | -0.16 | 0.10 | -0.36 – 0.03 | 0.097 |
| Mother achieved ≥ O level education | 0.27 | 0.29 | -0.31 – 0.84 | 0.359 | 0.22 | 0.26 | -0.29 – 0.73 | 0.396 |
| BMI of mother at enrollment | 0.07 | 0.04 | 0.00 – 0.14 | **0.047** | 0.13 | 0.03 | 0.07 – 0.19 | **<0.001** |
| Placental malaria present on histology | -0.45 | 0.31 | -1.06 – 0.16 | 0.148 | -0.73 | 0.28 | -1.27 – -0.18 | **0.009** |
| Female gender | 0.51 | 0.28 | -0.05 – 1.07 | 0.076 | 0.16 | 0.25 | -0.33 – 0.66 | 0.522 |
| Infant birth length (cm) | 0.15 | 0.06 | 0.04 – 0.26 | **0.009** | 0.12 | 0.05 | 0.02 – 0.22 | **0.020** |
| Age of infant (weeks) | 0.01 | 0.01 | -0.00 – 0.02 | 0.080 | -0.01 | 0.00 | -0.01 – -0.00 | **0.001** |

Length- and weight-for age Z scores defined using WHO Child Growth standards. SARS-CoV-2 infection during early pregnancy defined as IgM positive at enrollment in second trimester. SARS-CoV-2 infection during late pregnancy defined as seronegative in second trimester and seropositive at delivery. BMI=body mass index.

To further investigate the trajectory of infant growth by gestational SARS-CoV-2 infection status and potential synergy of placental malaria, we fit additional linear mixed effects models for length-and weight-for-age Z scores in infancy with interaction terms between SARS-CoV-2 infection and infant age, and between SARS-COV-2 infection and placental malaria (Figure S1). The interaction between early pregnancy SARS-CoV-2 infection and infant age had a small but significant positive association with the length-for-age Z score (Coef.=0.03, 95%CI 0.00-0.05, p=0.029), indicating that infants born to mothers infected with SARS-CoV-2 early in pregnancy had increasing length-for-age z scores over time compared with infants born to uninfected mothers, in contrast to a decrease or no change in infant length-for-age z scores over time in infants born to mothers with late pregnancy SARS-CoV-2 infection compared with no infection (Coef.=-0.02, 95%I −0.05-0.01, p=0.229; Figure 8K). This finding could suggest a delayed but time-limited effect of gestational SARS-COV-2 on infant height. Conversely, weight-for-age Z-scores decreased over time in infant born to SARS-CoV-2-infected mothers compared with uninfected mothers (Figure 8L). Interaction terms also revealed a potential synergistic negative effect of late pregnancy SARS-CoV-2 infection and placental malaria on both length- (Coef.=- 1.18, 95% CI −2.44-0.08, p=0.066) and weight-for-age (Coef.=-1.14, 95%CI −2.31-0.03, p=0.056) Z scores, but stratified analyses could not be performed due to low absolute numbers of women with both SARS-CoV-2 infection and placental malaria.

### Inferred variant SARS-CoV-2 infection and infant outcomes

Using the IgG binding ratios to different variants compared to Wuhan-Hu-1 to infer the viral variant responsible for a participant’s infection, we found that of the 168 with positive IgG at enrollment, 16 had predominant responses to Alpha variant Spike, 27 to Delta, 4 to Eta, and 1 to Gamma. Of 256 with positive IgG at delivery, 41 had predominant response to Alpha, 49 to Delta, and 15 to Gamma. Of the 320 participants with SARS-CoV-2 infection status in pregnancy able to be determined, 31 were presumed infected with Alpha variant, 48 with Delta, and 11 with Gamma. We note that, due to varied enrollment dates, viral variant waves occurred at different times during pregnancy for each participant. Comparing infants of Alpha versus Delta infected mothers, there was no difference in birth outcomes (Table S2). Lower length-for-age Z scores at 12 weeks of age were more common in infants whose mothers were infected with Delta, compared to Alpha variant in pregnancy (Figure 9).

**Figure 9.**
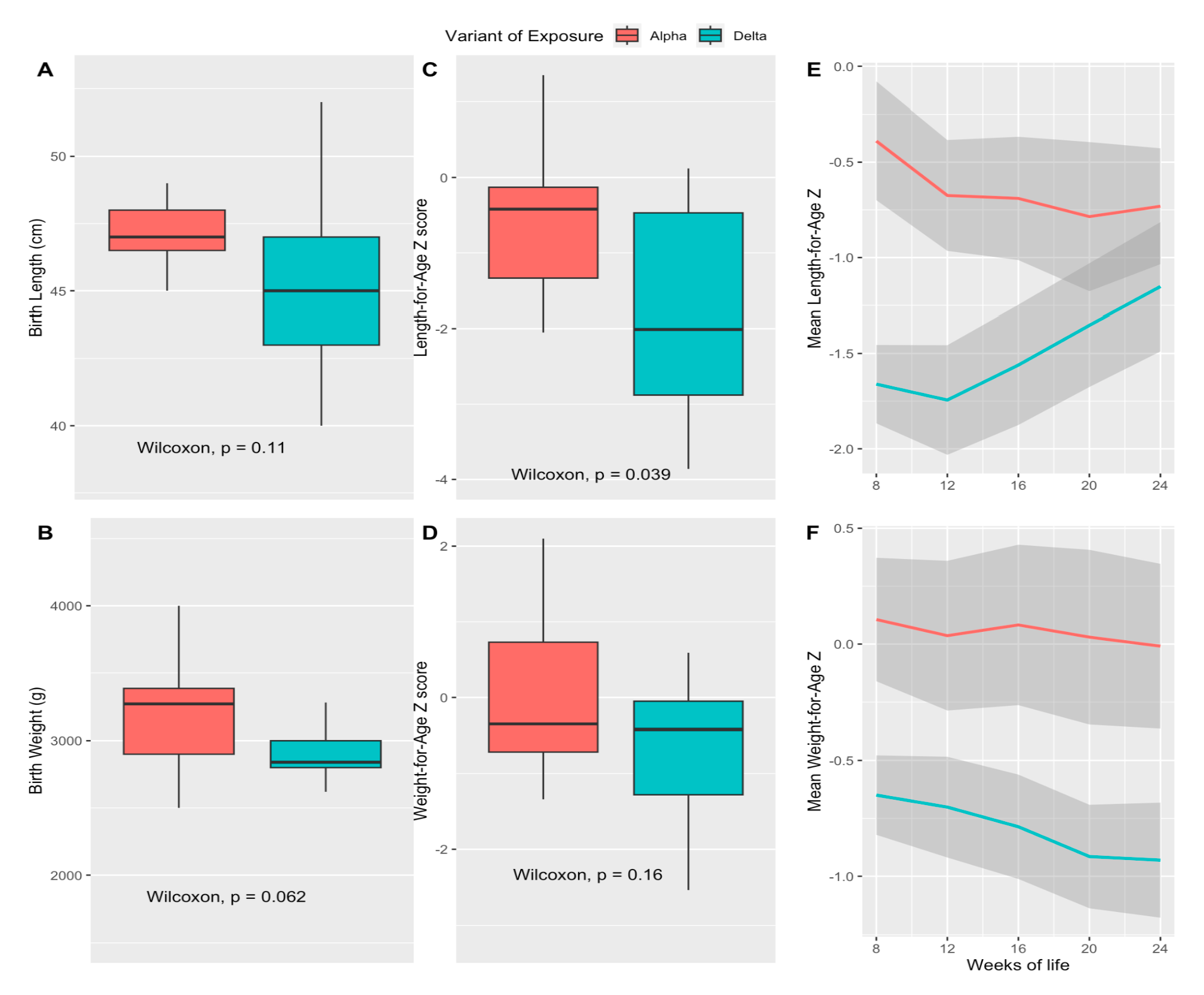
**Variant of exposure and association with infant growth through age 24 weeks** Length- and Weight-for-age Z scores determined from the WHO Child Growth reference standards. Boxplots revealing infant length at birth, infant weight at birth, length-for-age Z score at age 12 weeks, and length-for-age Z score at age 12 weeks comparing infants born to mothers exposed to SARS-CoV-2 Alpha versus Delta variant (A-D) and mean and 95% CIs (grey shaded region) of growth z scores from 8 to 24 weeks of life (E-F). Alpha=Exposed mother with Wuhan-Hu-1:alpha variant RBD IgG AU ratio<1 and lowest of all variant ratios. Delta=Exposed mother with Wuhan-Hu-1:delta variant RBD IgG AU ratio<1 and lowest of all variant ratios.

## Discussion

We observed high rates of SARS-CoV-2 exposure in this pregnancy cohort in eastern Uganda. We detected anti-Spike seropositivity rates similar to other serological surveys in Africa, showing increases in population seropositivity to SARS-CoV-2 from 12% in late 2020 to nearly 100% by early 2022 (4, 8). Some reports have described pre-pandemic cross-reactive antibodies to SARS-CoV-2 antigens in highly malaria infected populations (23), but we saw no significant difference in SARS-CoV-2 Spike binding IgG in plasma from pre-pandemic pregnant Ugandan women compared to pre-pandemic American influenza vaccination study participants. In contrast, pre-pandemic anti-nucleocapsid IgG and IgM, and anti-Spike IgM were higher in pregnant Ugandan women, and anti-RBD IgG was higher in the American controls. Taken together, these results do not support the hypothesis that pre-pandemic cross-reactive anti-coronavirus antibodies against SARS-CoV-2 Spike were a major protective factor for Africans.

Using precise internally-controlled comparisons of polyclonal plasma IgG binding to different viral variant antigens, we identified preferential binding to Wuhan-Hu-1-like antigens in individuals infected through early 2021, with a shift to more participants having preferential Alpha, Delta and other variant binding during later months of the pandemic. Thus, even if viral variant-specific nucleic acid testing is not available, serological testing can provide epidemiological data about viral variant exposures. Notably, the initial preferential binding of a patient’s plasma IgG to the likely variant infecting them was replaced over the course of months by broader binding to all viral variants in the panel of antigens tested. These data provide a population-level view of serological responses that are consistent with ongoing affinity maturation of IgG responses to SARS-CoV-2. Further analysis based on characterization of monoclonal antibodies derived from individual B cell clones detected over longitudinal sampling will be required to further characterize the mutational and affinity changes underlying these serological data. We note that responses to Omicron variants diverged from those to earlier antigenic variants in that participants infected during Omicron waves had IgG responses that bound Wuhan-Hu-1 antigens better than Omicron antigens; potential explanations for this lack of preferential binding to the likely infecting viral variant include the reported lower stability of Omicron Spike antigens, or prior asymptomatic infection with earlier variants that imprinted the humoral response with non-Omicron specificity (10, 24).

Despite the high rates of SARS-CoV-2 infection in this cohort, seroconversion was not associated with severe COVID-19 disease. This contrasts with evidence from large meta-analyses showing increased COVID-19 severity in pregnant women in many cohorts (25). Surges were associated with increased proportion of cough and upper respiratory infection-related visits, but not increased fever. Fever with COVID-19 may be less common in pregnant women than nonpregnant women, as has been reported in other African regions (11). SARS-CoV-2 infected participants had fewer headaches and fevers compared to uninfected participants, and no overall increase in cough or upper respiratory symptoms, suggesting that other respiratory infections or non-infectious environmental pulmonary exposures may have accounted for symptoms in many individuals. A subset of participants showed boosting of antibodies to endemic human coronaviruses NL63, HKU1, OC43, and 229E during the study period, suggesting that these viruses may have contributed to the respiratory symptoms reported. No participants developed severe respiratory disease consistent with severe COVID-19, although one participant experienced transverse myelitis, which has been reported in SARS-CoV-2 (26). Based on variant serology, this participant was likely infected with Alpha variant in the first trimester. She recovered with steroid treatment and had an uncomplicated birth.

We did not find any statistically significant associations between SARS-CoV-2 in pregnancy and birth outcomes including neonatal death, preterm birth, birth weight, birth length, or small for gestational age. One possible explanation for this may be that our sample size was insufficient to detect a difference in preterm birth or other birth outcomes. However, associations between gestational SARS-CoV-2 and preterm birth, pre-eclampsia, stillbirth, and low birth weight have been inconsistently reported in meta-analyses of observational clinical studies (25, 27). Further, symptomatic or severe COVID-19 is associated with higher risk of preterm birth compared with asymptomatic or mild cases (27), so the mild COVID-19 disease observed in our cohort may have been less likely to be associated with significantly increased adverse birth outcomes.

Despite the relatively small numbers of infants for whom follow up data was available, we noted differences in length and stunting in early infancy associated with maternal SARS-CoV-2 infection. To our knowledge, this is the first report of SARS-CoV-2 in pregnancy potentially affecting infant length (28). Many maternal infections are known to negatively impact fetal and infant weight including malaria (29), influenza (30), HIV (31) and Zika (32), but there are scant data on any particular gestational infection affecting length of offspring. Historical data suggests that *in utero* influenza exposure could negatively impact stature through at least eight years of age (33). In our data, stunting seemed greatest in infants born to mothers with positive IgM at enrollment, indicating early pregnancy infection, and in those with mothers infected with Delta variant, consistent with previous reports of Delta variant being more detrimental than other variants in pregnancy (34). Infections or inflammation early in pregnancy may disrupt limb development (established by eight weeks gestation) or growth *in utero*, contributing to height differences. On the other hand, infections in late pregnancy, particularly in the presence of placental malaria, may contribute more to weight differences, though this associations should be considered with caution due to the small number of women in this cohort with both SARS-CoV-2 infection and placental malaria. Further studies in larger cohorts are needed to better understand these potential associations.

Limitations to this study include reliance on retrospective clinical data from a trial of malaria therapies, reliance on serologic tests performed on banked specimens to determine SARS-CoV- 2 exposure given a lack of nucleic acid testing in real time, and a relatively small sample size to examine associations between gestational SARS-CoV-2 and clinical outcomes. Despite the inherent difficulties in retrospective serological data, these data characterize a large and clinically well-defined pregnancy cohort in East Africa during the first years of the SARS-CoV-2 pandemic. The sero-epidemiologic analysis carried out here with multiplexed MSD ECL assays is likely to be scalable to other populations to enable detailed tracking of viral variant infections, the serological responses to them, and clinical outcomes, even in settings where resource limitations preclude nucleic acid testing or direct virus detection.

## Data Availability

Underlying data and supporting analytic code for the article can be accessed upon request.

## Acknowledgements

We would like to thank all participants, research team members, ethics committee, funding agency, collaborating institutions, and patients, families, and caregivers for their valuable contributions to this clinical trial. Your support and dedication were essential to its successful completion.

