## Supplemental Materials for "High rates of SARS-CoV-2 infection in pregnant Ugandan women and association with stunting in infancy"

**Table S1. Linear mixed effects regression models to identify the effects of SARS-CoV-2 in pregnancy and other maternal and infant factors on length- and weight-for-age Z scores through 24 weeks of life with interaction terms**

| | Length-for-age Z score | | | | Weight-for-age Z score | | | |
|---|---|---|---|---|---|---|---|---|
| *Predictors* | *Estimates* | *std. Error* | *CI* | *p* | *Estimates* | *std. Error* | *CI* | *p* |
| **SARS-CoV-2 in pregnancy (Ref=Uninfected)** | | | | | | | | |
| **Early infection** | -1.75 | 0.46 | -2.66 – -0.85 | **<0.001** | -0.18 | 0.41 | -0.98 – 0.62 | 0.661 |
| **Late infection** | 0.56 | 0.50 | -0.42 – 1.55 | 0.263 | 1.02 | 0.43 | 0.17 – 1.87 | **0.019** |
| **Infant age (weeks)** | 0.00 | 0.01 | -0.02 – 0.02 | 0.761 | -0.00 | 0.00 | -0.01 – 0.01 | 0.861 |
| **Placental malaria** | -0.41 | 0.43 | -1.27 – 0.44 | 0.344 | -0.32 | 0.41 | -1.12 – 0.49 | 0.440 |
| **Gravidity** | -0.22 | 0.10 | -0.42 – -0.02 | **0.035** | -0.14 | 0.10 | -0.33 – 0.05 | 0.148 |
| **Mother achieved ≥ O level education** | 0.41 | 0.28 | -0.13 – 0.96 | 0.135 | 0.26 | 0.26 | -0.25 – 0.77 | 0.312 |
| **BMI of mother at enrollment** | 0.07 | 0.03 | 0.01 – 0.13 | **0.033** | 0.13 | 0.03 | 0.07 – 0.19 | **<0.001** |
| **Infant sex (female)** | 0.39 | 0.27 | -0.14 – 0.92 | 0.145 | 0.08 | 0.25 | -0.42 – 0.57 | 0.759 |
| **Infant birth length (cm)** | 0.13 | 0.05 | 0.02 – 0.23 | **0.019** | 0.11 | 0.05 | 0.01 – 0.21 | **0.026** |
| **Interaction terms:** | | | | | | | | |
| **Early infection x Infant age** | 0.03 | 0.01 | 0.00 – 0.05 | **0.029** | -0.01 | 0.01 | -0.02 – 0.00 | 0.067 |
| **Late infection x Infant age** | -0.02 | 0.02 | -0.05 – 0.01 | 0.229 | -0.02 | 0.01 | -0.03 – -0.01 | **0.007** |
| **Early infection x Placental malaria** | 0.72 | 0.59 | -0.43 – 1.87 | 0.219 | -0.30 | 0.55 | -1.39 – 0.78 | 0.586 |
| **Late infection x Placental malaria** | -1.18 | 0.64 | -2.44 – 0.08 | **0.066** | -1.14 | 0.60 | -2.31 – 0.03 | **0.056** |

Length- and weight-for age Z scores defined using WHO Child Growth standards. SARS-CoV-2 infection during early pregnancy defined as IgM positive at enrollment in second trimester. SARS-CoV-2 infection during late pregnancy defined as seronegative in second trimester and seropositive at delivery. BMI=body mass index.

### Table S2. Birth and infant outcomes by variant of exposure

| | Alpha (N=31) | Delta (N=48) | Total (N=79) | p value |
|---|---|---|---|---|
| **Spontaneous abortion** | 1 (3.2%) | 1 (2.1%) | 2 (2.5%) | 0.752 |
| **Stillbirth** | 0 (0.0%) | 1 (2.1%) | 1 (1.3%) | 0.421 |
| **Preterm birth** | 4 (13.3%) | 2 (4.3%) | 6 (7.9%) | 0.156 |
| **Low Birth Weight** | 1 (3.3%) | 3 (6.5%) | 4 (5.3%) | 0.543 |
| **Small for gestational age** | 4 (13.3%) | 6 (13.0%) | 10 (13.2%) | 0.971 |
| **Mother's WBC at delivery (cells/µl)** | 10840 (8935, 13835) | 11350 (9110, 13050) | 11200 (9013, 13398) | 0.713 |
| **Female gender** | 12 (40.0%) | 22 (46.8%) | 34 (44.2%) | 0.557 |
| **Birth Weight (g)** | 3080 (2945, 3325) | 2950 (2800, 3268) | 3000 (2800, 3280) | 0.310 |
| **Birth length (cm)** | 47 (46, 48) | 47 (44-48) | 47 (45-48) | 0.482 |
| **Placental malaria** | 9 (36.0%) | 14 (37.8%) | 23 (37.1%) | 0.883 |
| **Height at 12 weeks (cm)** | 59 (58-60) | 56 (55-59) | 58 (55-59) | 0.037 |
| **Weight at 12 weeks (kg)** | 5.69 (5.40, 6.76) | 5.53 (5.00, 6.02) | 5.55 (5.28, 6.14) | 0.088 |
| **Length-for-age Z score at 12 weeks** | -0.42 (-1.34, -0.13) | -2.01 (-2.88, -0.47) | -1.34 (-2.31, -0.360 | 0.037 |
| **Weight-for-age Z score at 12 weeks** | -0.35 (-0.72, 0.73) | -0.42 (-1.28, -0.05) | -0.39 (-1.02, 0.10) | 0.159 |
| **Stunting at 12 weeks** | 1 (9.1%) | 11 (52.4%) | 12 (37.5%) | 0.016 |
| **Underweight at 12 weeks** | 0 (0.0%) | 2 (9.5%) | 2 (6.2%) | 0.290 |

IQR=Interquartile range. Length- and Weight-for-age Z scores determined from WHO reference standards. P values are reported for chi-square tests of proportions for categorical variables [displayed as n (%)], and Mann-Whitney U tests for continuous variables [displayed as median, (IQR)].

**Figure S1. Comparison of Spike antibody levels as measured by Luminex vs MSD**

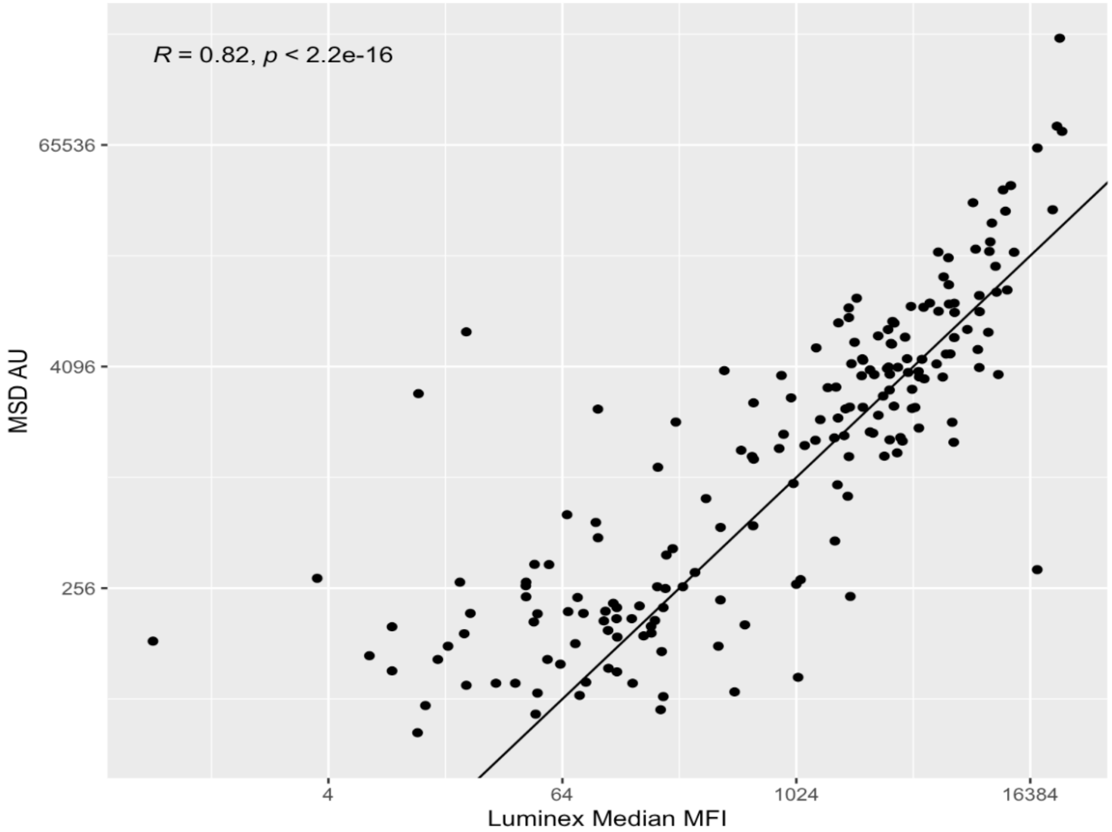


Correlation of Luminex SARS-CoV-2 Spike IgG median MFI with SARS-CoV-2 Wuhan-Hu-1 Spike IgG AU as measured on MSD platform.
